# APOE-stratified genome-wide association study suggests potential novel genes for late-onset Alzheimer’s disease in East-Asian descent

**DOI:** 10.1101/2020.07.02.20145557

**Authors:** Sarang Kang, Jungsoo Gim, Tamil Iniyan Gunasekaran, Kyu Yeong Choi, Jang Jae Lee, Eun Hyun Seo, Pan-Woo Ko, Ji Yeon Chung, Seong-Min Choi, Young Min Lee, Jee Hyang Jeong, Kyung Won Park, Min Kyung Song, Ho-Won Lee, Ki Woong Kim, Seong Hye Choi, Dong Young Lee, Sang Yun Kim, Hoowon Kim, Byeong C. Kim, Takeshi Ikeuchi, Kun Ho Lee

**Author notes:** Correspondence: Correspondence to: Jungsoo Gim, PhD, Department of Biomedical Science, Chosun University, Gwangju 61452, Republic of Korea, and Kun Ho Lee, PhD, Department of Biomedical Science, Chosun University, Gwangju 61452, Republic of Korea. Note: These authors contributed equally to this work (Sarang Kang, Jungsoo Gim).

## Abstract

In this study, we report two new possible susceptible genes for late-onset Alzheimer’s disease identified through an APOE-stratified genome-wide association analysis (GWAS) using East Asian samples. In the discovery phase, we performed a GWAS of Alzheimer’s disease in 2,291 Korean seniors from the Gwangju Alzheimer’s and Related Dementias (GARD) cohort study. A successive replication analysis with a Japanese sample of size 1,956 suggested three novel susceptible SNPs in two genes: *LRIG1* and *CACNA1A*. This study demonstrates that the discovery of AD-associated variants can be accomplished in non-European ethnic groups with a more homogeneous genetic background using samples comprising fewer subjects.

## INTRODUCTION

A genetic component has consistently been identified as an important antecedent factor in Alzheimer’s disease (AD), and Apolipoprotein E (*APOE*) is the most prevalent risk factor of the disease [1, 2]. The effect of the *APOE* genotype, however, varies greatly among populations [3]. One noteworthy example is that the e4 allele, the risk form of *APOE*, is virtually absent among Arabs living in the northern Israeli community where the prevalence of dementia (in spite of unspecified pathology) is roughly double that in European ancestry [4].

Since the identification of the only locus in *APOE* with a sample consisting of 1,086 individuals in 2007 [5], genome-wide association studies (GWASs) have increased the sample size to improve the statistical power to identify the missing causal variants for late-onset AD (LOAD) [6, 7]. The latest and largest GWAS expands to include 1,126,563 cases and controls (including 364,859 proxy) [Douglas P. Wightman, et al., 2020, medRxiv: https://doi.org/10.1101/2020.11.20.20235275]. The variants and associated genes from those GWASs were identified mostly in individuals of European ancestry and found enriched in various biological pathways, including immune responses and endocytosis, suggesting genetic heterogeneity or the complex nature of the LOAD.

Genetic studies in diverse populations have increased our understanding of the genetic architecture of LOAD [8-12]. Studies of non-European ancestry populations might have a chance of being used to discover variants that are rare or absent, thus showing a smaller effect size in European ancestry [13]. Notably, the discovery of such AD-susceptible loci, displaying allelic heterogeneity among diverse populations, has been accomplished with non-European ancestry groups of a more homogeneous genetic background using samples comprising several thousand or much fewer subjects [10, 11].

In the present study, we leverage the genetic architecture of Koreans, a homogeneous East Asian population maintaining a distinct genetic profile with a high prevalence of AD among seniors (65 years and older), to discover AD-associated variants. For this purpose, we first performed GWAS and post-hoc GWAS with APOE stratified in Koreans using the Korea Biobank Array (referred to as Korean Chip) designed for Koreans [14] and replicated the findings by combining genotype level data from another East Asian population [11].

## MATERIALS AND METHODS

### Study Sample

We analyzed genotype level data of 2,291 individuals (1,119 AD cases and 1,172 controls) for the discovery stage, and 1,956 individuals (980 AD cases and 976 controls) for the replication stage (**Table 1**). Participants for the discovery stage were sampled from the Gwangju Alzheimer’s & Related Dementias (GARD) Cohort Research Center, a longitudinal single-center study designed to develop clinical, imaging, genetic, and other biomarkers for early detection and tracking of dementia (mostly AD) from local senior citizens aged 60 years or older in Gwangju, South Korea. The study protocol was approved by the Institutional Review Board of Chosun University Hospital, Korea (CHOSUN 2013-12-018-070). All volunteers or authorized guardians for cognitively impaired individuals gave written informed consent before participation. Out of 16,002 participants in the GARD cohort, 2,291 (1,119 AD dementia and 1,172 cognitive normal) were chosen to maximize contrast between AD cases and cognitive normal controls for the GWAS. All cases were at least 60 years old and met NINCDS-ADRDA criteria for AD [15], while cognitively healthy controls were at least 70 years old and either judged to be cognitively normal or did not meet pathological criteria (**Supplementary Material**).

**Table 1.** Summary of demographic information for each group in GWAS analyses.

|  | Discovery (n=2,291) |  | Replication (n=1,956) |  |
| --- | --- | --- | --- | --- |
|  | CN | AD | CN | AD |
| All samples, n | 1,172 | 1,119 | 976 | 980 |
| Female, n (%) | 661 (56.4) | 715 (63.9) | 564 (57.8) | 702 (71.6) |
| Age, m (s.d) | 76.03 (8.9) | 74.60 (8.9) | 76.94 (5.9) | 72.99 (4.3) |
| APOE e4 non-carrier, n | 976 | 621 | 815 | 435 |
| Female, n (%) | 544 (55.7) | 393 (63.3) | 470 (57.7) | 31 (71.3) |
| Age, m (s.d) | 76.10 (8.9) | 75.43 (9.1) | 77.11 (5.9) | 73.42 (4.3) |
| APOE e4 carrier | 196 | 498 | 161 | 545 |
| Female, n (%) | 117 (59.7) | 322 (64.7) | 94 (58.4) | 392 (68.3) |
| Age, m (s.d) | 95.45 (8.9) | 73.32 (9.0) | 76.06 (5.9) | 72.65 (4.3) |

### Genotyping, Imputation, and Gene Mapping

We applied extensive quality control measures used in the standard protocol [16] and excluded subjects with a low genotyping success rate (95%), heterozygosity outliers and those that exhibited cryptic relatedness. We assessed study population heterogeneity by means of a principal component analysis (PCA) and multidimensional scaling (MDS) and removed those that did not meet the criteria before proceeding with further analyses. We also excluded those with SNPs with a minor allele frequency <1%, a genotyping success rate <95%, or a deviation of the genotype distribution from Hardy-Weinberg equilibrium in the control group (P < 1×10^−6^). To increase the genotyping coverage between the two datasets, we imputed missing genotypes and SNPs using pre-phased reference haplotypes from the Haplotype Reference Consortium (HRC) panel version 1.1 for each dataset. We subjected all imputed SNPs to the same quality criteria described above and added the requirement of imputation quality with an info score >0.5, leaving 35,685,761 and 39,044,005 SNPs for discovery and replication analysis, respectively. Functional mapping of SNPs to genes was conducted using a gene annotation file from Affymetrix [14] and the SNPnexus web tool [17].

### Statistical analysis

A set of GWAS analyses for AD status was performed in three models of a two-stage framework (discovery and replication): 1) all samples; 2) APOE e4 carriers; and 3) APOE e4 non-carriers. The GWAS was performed separately in each model for both the discovery and replication stages. The association of genotype dosages of each SNP in the additive component with the AD case-control status was estimated using logistic regression analysis adjusting for age, sex, and first four principal components using PLINK and EIGENSTRAT software [18-20]. Only the SNPs that attained a suggestively significant association level (P < 5×10^−5^) were tested in the replication stage using a Japanese dataset. A the total of 4,247 subjects was assembled for meta-analysis and SNP effect estimates and their SEs were combined by a fixed effect model with inverse variance weighted method using the METAL software [21]. SNP heritability was estimated using the GCTA software [22].

## RESULTS

### Genome-wide inference for AD status

For discovery, we first conducted GWA analysis with the GARD sample, which included 2,291 subjects and identified 54 genome-wide significant (5E-08) SNPs across three genes (genomic inflation = 1.03), and all those genes (*APOE, PVRL2*, and *TOMM40*) were found to be associated with LOAD in previous studies (**Table 2**). We estimated the liability-scale SNP heritability, excluding those loci, and a relatively large estimate (0.566±0.152) was obtained. In addition, a portion of SNPs in other previously known genes, such as *ABCA7* and *BIN1*, was replicated at a genome-wide suggestive level (5E-05). These results suggest that additional common variants with smaller effects (or possibly rarer variants with larger effects) yet to be found might be identified through a post-hoc analysis with a less stringent statistical significance.

**Table 2.** Top-ranked genome-wide association results in the Korean discovery sample (P < 5×10^−5^) and their replication in Japanese and meta-analyses.

| Gene | Chr | Lead SNP (type)* | BP | m/M | MAF | Discovery (1,172/1,119) |  | Replication (976/980) |  | Meta-analysis |  | Previously reported GWAS |  |  | Alt Allele Frq<sup>†</sup> |
| --- | --- | --- | --- | --- | --- | --- | --- | --- | --- | --- | --- | --- | --- | --- | --- |
|  |  |  |  |  |  | OR (95% CI) | P | OR (95% CI) | P | OR | P | GENE (SNP) | P | ref |  |
| ALL samples |  |  |  |  |  |  |  |  |  |  |  |  |  |  |  |
| APOE | 19 | rs429358 (m) | 45411941 | C/T | 0.169 | 3.637 (3.03-4.37) | 3.74×10<sup>-43</sup> | 2.608 (2.04-3.33) | 1.89×10<sup>-14</sup> | 3.227 | 5.90×10<sup>-55</sup> | rs429358 | 1.4×10<sup>-546</sup> | [9] | 0.036 |
| TOMM40 | 19 | rs10119 (3') | 45406673 | A/G | 0.180 | 3.378 (2.83-4.03) | 9.58×10<sup>-42</sup> | 2.230 (1.88-2.65) | 7.49×10<sup>-20</sup> | 2.731 | 1.59×10<sup>-57</sup> | rs10119 | 1.2×10<sup>-342</sup> | [36] | 0.279 |
| PVRL2 | 19 | rs12972156 (i) | 45387459 | G/C | 0.151 | 2.942 (2.45-3.54) | 1.52×10<sup>-30</sup> | 2.656 (2.06-3.43) | 5.42×10<sup>-14</sup> | 2.841 | 8.30×10<sup>-43</sup> | PVRL2 (rs412776) | 2.2×10<sup>-21</sup> | [9] | 0.001 |
| APOC1 | 19 | rs10414043 (g) | 45415713 | A/G | 0.143 | 3.082 (2.55-3.73) | 3.92×10<sup>-31</sup> | 2.629 (2.02-3.43) | 8.76×10<sup>-13</sup> | 2.920 | 4.29×10<sup>-42</sup> | rs10414043 | 1.2×10<sup>-522</sup> | [36] | 0.109 |
| APOE non-carrier |  |  |  |  |  |  |  |  |  |  |  |  |  |  |  |
| CACNA1A | 19 | rs189753894 (i) | 13624489 | A/C | 0.082 | 1.726 (1.34-2.23) | 2.68×10<sup>-5</sup> | 1.900 (1.35-2.67) | 2.22×10<sup>-4</sup> | 1.787 | **2.49×10<sup>-8</sup>** | - | - | - | 0.007 |
| LRIG1 | 3 | rs2280575 (i) | 66542863 | G/A | 0.092 | 0.539 (0.41-0.71) | 1.15×10<sup>-5</sup> | 0.551 (0.40-0.76) | 3.46×10<sup>-4</sup> | 0.544 | **1.51×10<sup>-8</sup>** | - | - | - | 0.267 |
| APOE e4 carrier |  |  |  |  |  |  |  |  |  |  |  |  |  |  |  |
| - | - | - | - | - | - | - | - | - | - | - | - | - | - | - | - |

\*m: missense, s: synonymous, i: intronic, g: intergenic, 3': 3' UTR; <sup>†</sup>SNP alternative allele frequencies of Europeans reported in the allele frequency aggregator (ALFA) [37]

We carried out *APOE*-stratified GWA analysis using subgroups of *APOE* e4 carrying status. A total of 219 variants (annotated in 61 genes) and 306 variants (in 82 genes) were respectively identified as e4 carriers (genomic inflation = 1.02) and non-carriers (genomic inflation = 1.03) with a suggestive level of 5E-05. Loosening statistical significance might increase false findings; thus, we included an independent cohort dataset for replication. For valid novel discovery, we performed a meta-analysis with the replication dataset, filtered with previous GWAS signals, resulting in examination of 7 out of 61 and 15 out of 82 genes for e4 carriers and non-carriers, respectively. Considering the effect direction and statistical significance of the variants in both the discovery and replication datasets, *CACNA1A* (OR = 1.726, lead SNP rs189753894) and *LRIG1* (OR = 0.539, lead SNP rs2280575) remained in the association result for further functional annotation (**Table 2**).

### Functional annotation

We followed up the association results of two novel genes, *CACNA1A* and *LRIG1*, by carrying out functional annotation using web-based platforms from the Human Protein Atlas (HPA) [23], Genotype-Tissue Expression (GTEx) [24], and Functional Annotation of Mammalian Genomes 5 (FANTOM5) [25]. *CACNA1A*, a risk gene candidate, is on chromosome 19. However, it is not likely to be influenced by the *APOE* signal because *CACNA1A* is far from the *APOE* locus (more than 32Mb) and is identified with *APOE* non-carriers. We also confirmed its independence with all subjects by conditional analysis with *APOE*\*4 (rs429358). Tissue specificity analysis with GTEx, FANTOM5, and HPA showed that *CACNA1A* is expressed in brain tissues, particularly in the cerebellum at both the transcription and protein levels (**Supplementary Fig. 1A, Fig. 1A**). The *CACNA1A* gene plays an important role in neuronal cell death. It codes subunits of neuronal calcium channels, which are involved in the neuronal cell death and toxicity of amyloid beta [26, 27].

**Figure 1.**
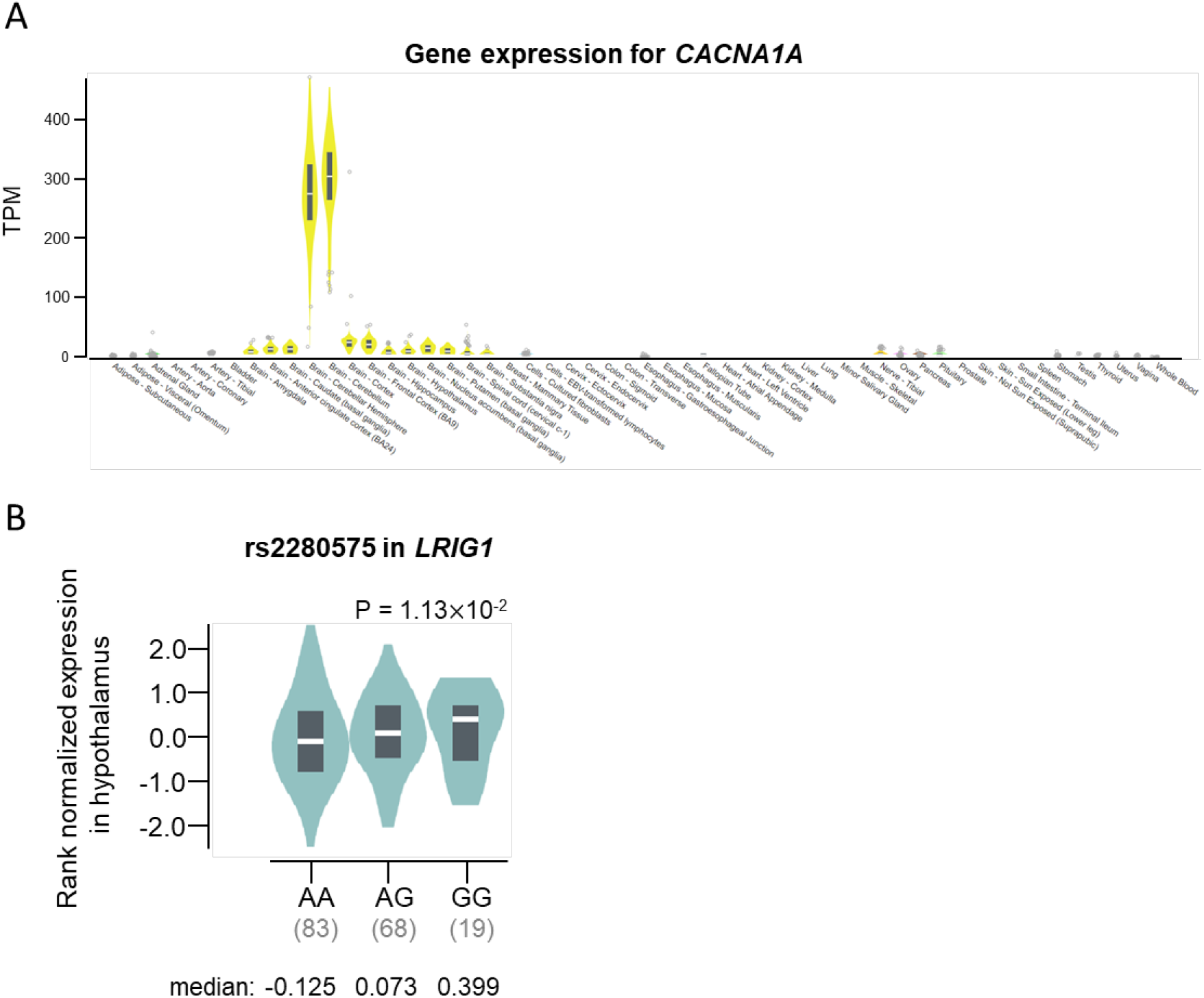
Gene expression of *CACNA1A* across different tissues (B) and expression quantitative trait loci (eQTL) analysis of rs2280575 in *LRIG1* (A). (A) The *CACNA1A* is expressed at a high level in the cerebellum (median TPM: 304.8) and in the cerebellar hemisphere (median TPM: 275.0). (B) Violin plot showing the effect of the eQTL rs2280575 on *LRIG1* expression in hypothalamus (P = 1.13×10^−2^). (Image available from https://gtexportal.org/home/gene/CACNA1A).

Expression levels of *LRIG1* were high in the cerebral cortex at 74.1 protein-coding transcripts per million (pTPM) in HPA, 16.0 pTPM reported by GTEx (**Supplementary Fig. 1B**). In the FANTOM5 dataset, the RNA expression levels of *LRIG1* were higher in the brain tissues than in other tissues, especially for the hippocampal formation, which was 179.9 scaled tags per million. eQTL analysis with lead SNP rs2280575 in *LRIG1* showed significant results in the hypothalamus (**Fig. 1B**). The association between *LRIG1* and the hippocampus was evaluated in *in vitro* and *in vivo* analyses in a previous study [28].

## DISCUSSION

Here, we conducted GWA analysis with a Korean sample to identify AD-associated loci. We validated the association between AD status and *APOE* locus in chromosome 19 with genome-wide significance. To mask the effect of *APOE* and identify novel candidate loci in East Asians, we carried out an APOE4 stratified GWAS with two independent East Asian samples and identified *CACAN1A* (lead SNP rs189753894) and *LRIG1* (lead SNP rs2280575) among *APOE4* non-carriers.

From the functional annotation and literature survey, we found that the two genes play an essential role in neurobiological function. *CACNA1A*, a subunit of the neuronal calcium channel, is predominantly and specifically expressed in brain tissue and was found to be involved in neuronal cell death and toxicity of amyloid beta, suggesting a plausible risk gene for AD. The genetic function of *LRIG1* has been shown to be involved in the dendritic formation of neurons, as well as in affecting the neural cell function of the hippocampus. Abnormal expression and dysfunction of *LRIG1* lead to dendritic abnormalities, which are involved in morphogenesis of hippocampal dendrites by brain-derived neurotrophic factor (BDNF) signaling. BDNF plays a major role in the growth, development and survival of neurons, and is involved in synaptic plasticity and synaptogenesis for learning and memory in the adult brain [29-31]. BDNF showed low gene expression levels in patients with AD [32]. Several studies have shown that high expression levels of BDNF could slow down cognitive decline in the elderly and in patients with AD [29, 33-35].

There are limitations to this study that need to be considered. The sample size in this study is relatively smaller than those in other AD GWA studies. However, the sample used in this analysis was a representative sample with well-defined cases and controls: subjects to maximize contrast were chosen, and any subjects with ambiguous diagnosis were removed from the further analysis. Although the previous studies about the function of *CACNA1A* and *LRIG1* suggest their role in the development of the LOAD, a conclusion whether each gene has an impact on the progression of dementia or protecting neurons from degeneration requires further biological validation with an AD model system.

To conclude from the findings made in this work, we validated a portion of previously reported LOAD-associated genes, identified from the mostly Caucasian subjects, using the Koreans. We also identified and replicated East Asian-specific novel loci in *CACNA1A* and *LRIG1* through a post-hoc GWAS with APOE4 stratified. The finding in this work might provide an improved understanding of complex genetic signature of LOAD.

## Data Availability

The data that support the findings of this study are available on request from the corresponding author, [K.H.L]. The data are not publicly available due to [restrictions e.g. their containing information that could compromise the privacy of research participants].

## Acknowledgments

This research was supported by the Original Technology Research Program for Brain Science of the National Research Foundation (NRF-2014M3C7A1046041 and NRF-2014M3C7A1046042), and KBRI basic research program (20-BR-03-02) funded by Koera Ministry of Science and ICT. This study was also supported by Japan Agency for Medical Research and Development (AMED) (JP18kk0205009).

## IRB

The study protocol was approved by the Institutional Review Board of Chosun University Hospital, Korea (CHOSUN 2013-12-018-070). All volunteers or authorized guardians for cognitively impaired individuals gave written informed consent before participation.

