## Supplementary Material for "APOE-stratified genome-wide association study suggests potential novel genes for late-onset Alzheimer’s disease in East-Asian descent"

#### *GARD Cohort*

The GARD (Gwangju Alzheimer's & Related Dementias) cohort is a group that has been tracked since 2010 after recruiting local senior citizens aged 60 years or older for research on Alzheimer's disease and dementia. Most of the subjects were recruited at the Early Dementia Examination Center in Gwangju Bitgoeul Senior Health Town to verify basic personal information, academic background, medical history, family history, and drug use, and to conduct biometric (EEG, PPG, impedance) tests and K-MMSE (Korean Mini-Mental State Examination) tests to determine whether the subject suffered from dementia. Among subjects whose K-MMSE assessment had been completed, those whose MMSE score >27 were selected, and the number of subjects recruited was currently 16,002. An additional thorough clinical examination was conducted for those who wished to have one conducted and those who agreed to the study after conducting the basic examination. Clinical evaluation was conducted through brain imaging information of subjects using the Seoul Neuropsychological Screening Battery (SNSB) [1] test and 3T MRI (Skyra, Siemens) single scanner. $^{18}$F-Florbetaben (FBB) beta-amyloid positron emission tomography (PET) was performed on some of the subjects. All diagnoses were evaluated by dementia specialists in neurology and psychiatry at Chosun University Hospital and Chonnam National University Hospital. Blood tests (complete blood count, cholesterol, blood sugar, thyroid function test), CSF testing were carried out on the diagnosed subjects. Genomic DNA was extracted from peripheral blood leukocytes that were isolated from whole blood collected in an EDTA tube. Cognitively normal (CN) subjects had no evidence of neurological disorders and impairment in cognitive function or activities of daily living. Excluded subjects included those who had less than three years of education, had a history of brain disease and poor mental health, were taking related drugs, were consuming high levels of alcohol or had depression. The clinical diagnosis of probable AD was determined using the NINCDS/ADRDA criteria [2]. A portion of AD blood species was from other major hospitals in Korea, including Seoul National University Hospital, Kyungpook National University Hospital, Inha University Hospital, and Pusan National University Hospital, and the use of species was approved by the IRB of Chosun University Hospital.

### LEGEND TO FIGURES AND TABLES

**Supplementary Figure 1.** The RNA expression and distribution of CACNA1A (A) and LRIG1 (B) in brain region (yellow bar) among normal human tissues. (A) Data from HPA dataset showing highest mRNA expression of the CACNA1A gene in the cerebral cortex at 18.2 protein-coding transcripts per million (pTPM). The expression of the CACNA1A gene was highest in the cerebellum at 394.1 pTPM in GTEx, 453.4 scaled-tags per million reported by FANTOM5. (B) The expression levels of LRIG1 were high in the cerebral cortex at 74.1 pTPM in HPA, 16.0 pTPM reported by GTEx. Data from FANTOM5 dataset showing highest mRNA expression of the LRIG1 gene in hippocampal formation at 179.9 scaled-tags per million. (Imgage available from https://www.proteinatlas.org/ENSG00000141837-CACNA1A/tissue and https://www.proteinatlas.org/ENSG00000144749-LRIG1/tissue)

**Supplementary Figure 1.**

A

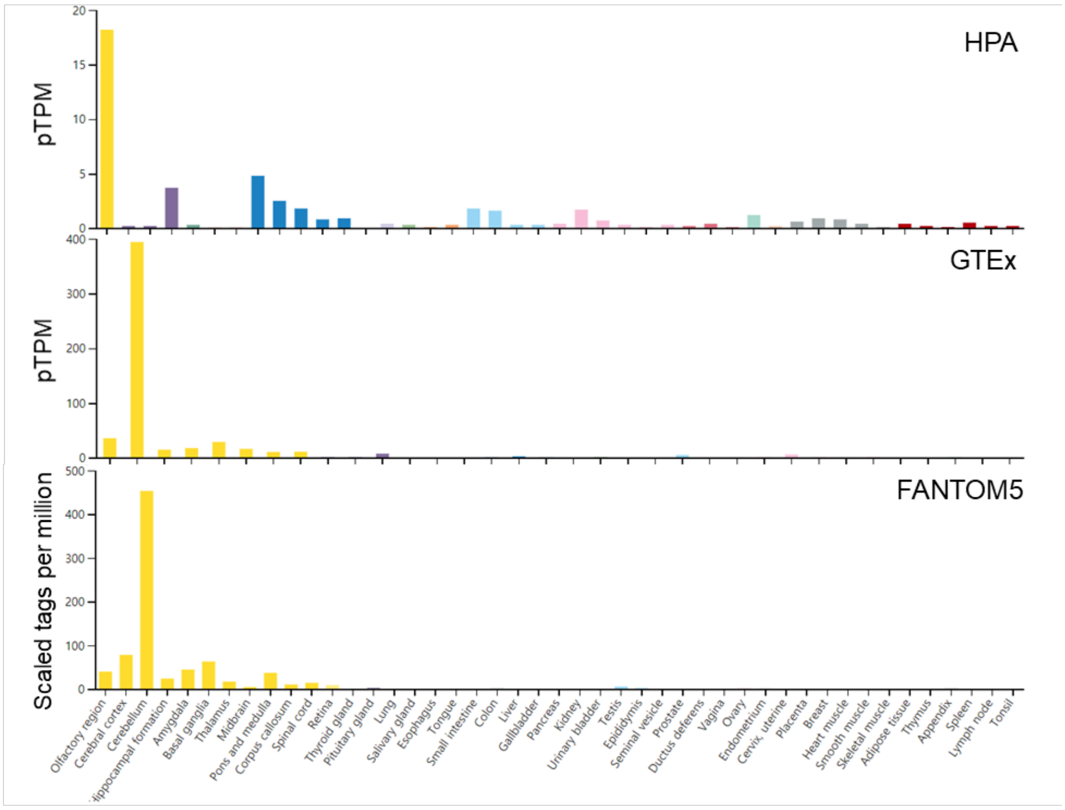


B

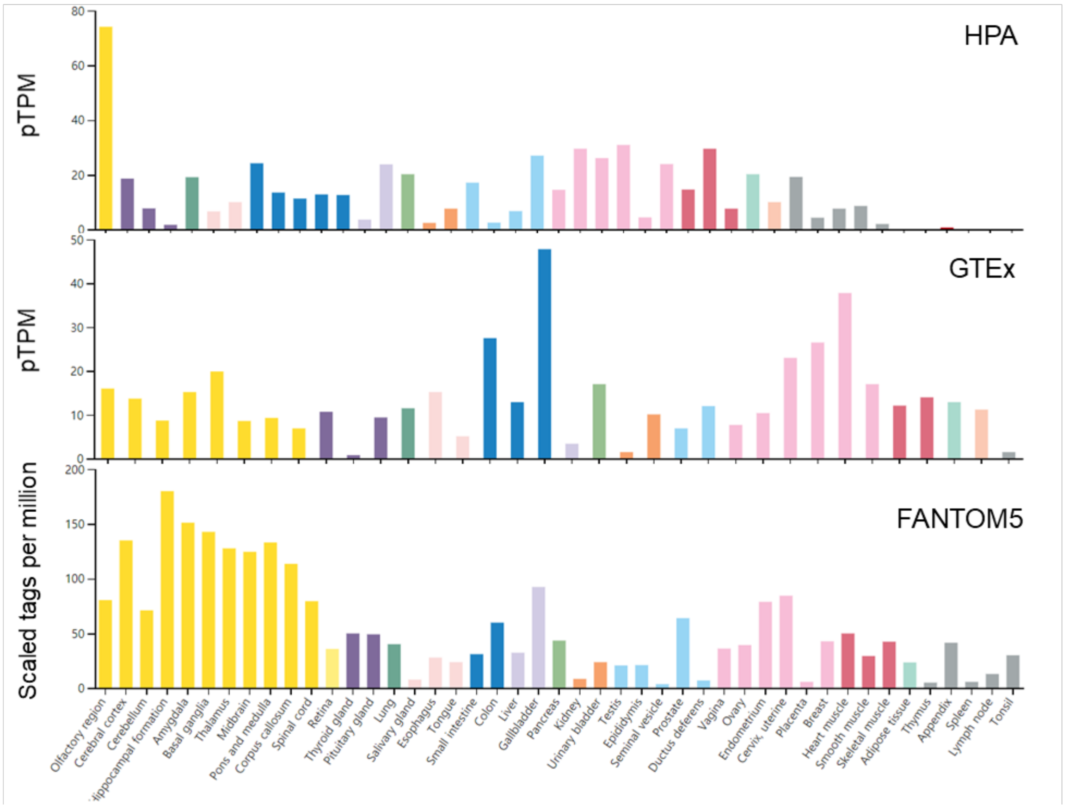
